# Conversational Artificial Intelligence for Translational Precision Medicine: Integrating Social Determinants of Health, Genomics, and Clinical Data with AI-HOPE-PM

**DOI:** 10.1101/2025.03.28.25324864

**Authors:** Ei-Wen Yang, Brigette Waldrup, Enrique Velazquez-Villarreal

## Abstract

Introduction: Achieving equity in translational precision medicine requires the integration of genomic, clinical, and social determinants of health (SDoH) data to uncover disease mechanisms, personalize treatment, and reduce health disparities. Yet, existing bioinformatics tools are often hindered by fragmented data structures, steep technical barriers, and limited capacity to incorporate SDoH variables-challenges that disproportionately affect underserved populations. To address this, we developed AI-HOPE-PM (Artificial Intelligence agent for High-Optimization and Precision mEdicine in Population Metrics), a conversational AI platform that allows users to conduct multi-dimensional cancer analyses through natural language interaction. By unifying large-scale clinical, genomic, and SDoH data within a dynamic and accessible interface, AI-HOPE-PM lowers the barrier to integrative research and supports inclusive, hypothesis-driven investigation. Methods: AI-HOPE-PM leverages large language models (LLMs), structured natural language processing, retrieval-augmented generation (RAG), and an internal Python-based workflow engine to automate data ingestion, filtering, cohort stratification, and statistical analysis. The platform operates on harmonized datasets from TCGA, cBioPortal, and AACR GENIE, enriched with simulated SDoH variables such as financial strain, food insecurity, and healthcare access. Free-text queries (e.g., Compare survival outcomes in CRC patients with TP53 mutations and limited access to care) are parsed into executable scripts aligned with biomedical ontologies. The system performs survival modeling, odds ratio testing, and case-control comparisons, generating interpretable visualizations and narrative reports in real time. Benchmarking against platforms like cBioPortal and UCSC Xena demonstrated 92.5% query interpretation accuracy and efficient performance across both CPU and GPU cloud environments. Results: AI-HOPE-PM successfully translated diverse user queries into real-time, executable analyses across colorectal cancer (CRC) datasets, enabling integration of clinical, genomic, and SDoH data. In one case study, the platform identified significantly worse survival in FOLFOX-treated CRC patients with TP53 mutations experiencing financial strain (p = 0.0481). Another analysis revealed poorer progression-free survival in APC wild-type patients with good healthcare access (p = 0.0233). Additional findings highlighted the influence of social support (p = 0.0220), food insecurity (p = 0.0162), and health literacy on outcomes and treatment access. Odds ratio analyses revealed disparities in chemotherapy exposure (OR = 0.356 for food-insecure patients) and KRAS mutation prevalence by sex and literacy status. AI-HOPE-PM also surfaced racial and ethnic differences in progression-free survival, emphasizing the importance of SDoH integration in population-level cancer research. All analyses were completed in under one minute, significantly reducing manual workload and improving scalability. Conclusions: AI-HOPE-PM marks a significant leap forward in the field of precision oncology by uniting clinical, genomic, and SDoH data within a single, conversational AI framework. Instead of relying on traditional, code-heavy approaches, the platform enables users to perform complex, multi-layered analyses through simple natural language interactions. This functionality not only democratizes access to integrative cancer research but also enhances the ability to uncover disparities in outcomes linked to genetic, clinical, and social variables. By contextualizing molecular insights within real-world social environments, AI-HOPE-PM delivers a more comprehensive understanding of cancer biology and care inequities. Its high performance, interpretability, and scalability position it as a powerful tool for accelerating hypothesis generation, guiding biomarker discovery, and informing equity-driven treatment strategies. As a flexible and user-centered platform, AI-HOPE-PM lays the groundwork for a new paradigm in AI-assisted, health equity-focused translational research.

## Introduction

Healthcare is being transformed by comprehensive precision medicine, which personalizes treatment based on individual differences in genetics, environment, and lifestyle [1,2]. Alongside this shift, there is growing recognition of the critical role social determinants of health (SDoH) play in shaping disease outcomes and access to care [2–5]. To advance both scientific discovery and health equity, integrating clinical, genomic, and SDoH data is imperative for uncovering disease mechanisms, enhancing treatment effectiveness, and reducing disparities—especially among underserved populations. However, several challenges remain: data silos, the need for specialized expertise in multi-omics analysis, and the underrepresentation of diverse populations in existing datasets all continue to hinder the equitable realization of precision medicine [6–9].

The complexity of cancer research workflows demands seamless integration of molecular profiles, clinical metadata, and population-level variables such as race, ethnicity, income, health literacy, and access to care. Although web-based tools like cBioPortal [10] and UALCAN [11] offer structured platforms for querying public cancer datasets such as The Cancer Genome Atlas (TCGA) [12], they operate within predefined analytical frameworks and require users to manually conduct multi-step filtering, stratification, and statistical interpretation [13, 14]. These limitations restrict the flexibility needed to explore hypothesis-driven, context-specific research questions— especially those involving SDoH variables critical for addressing health equity.

Meanwhile, emerging AI-based tools like CellAgent [15] and AutoBA [16] have begun to explore the potential of large language models (LLMs) in bioinformatics workflows [17–20]. However, these systems often focus solely on genomic data and lack the capacity to simultaneously integrate clinical and SDoH variables, thereby limiting their utility in advancing equitable biomedical research.

Motivated by these gaps, we introduced AI-HOPE-PM (Artificial Intelligence agent for High-Optimization and Precision mEdicine in Population Metrics), a novel LLM-powered conversational agent designed to democratize access to integrative bioinformatics analysis. AI-HOPE-PM allows users—regardless of technical background—to conduct robust, multi-dimensional cancer research using natural language queries. As illustrated in Figure 1, the platform employs natural language processing (NLP), retrieval-augmented generation (RAG), and Python-based bioinformatics pipelines to translate user queries into reproducible and explainable analyses. This includes case-control comparisons, survival modeling, and stratified multi-omics analysis—all without requiring code or manual data preprocessing.

**Figure 1.**
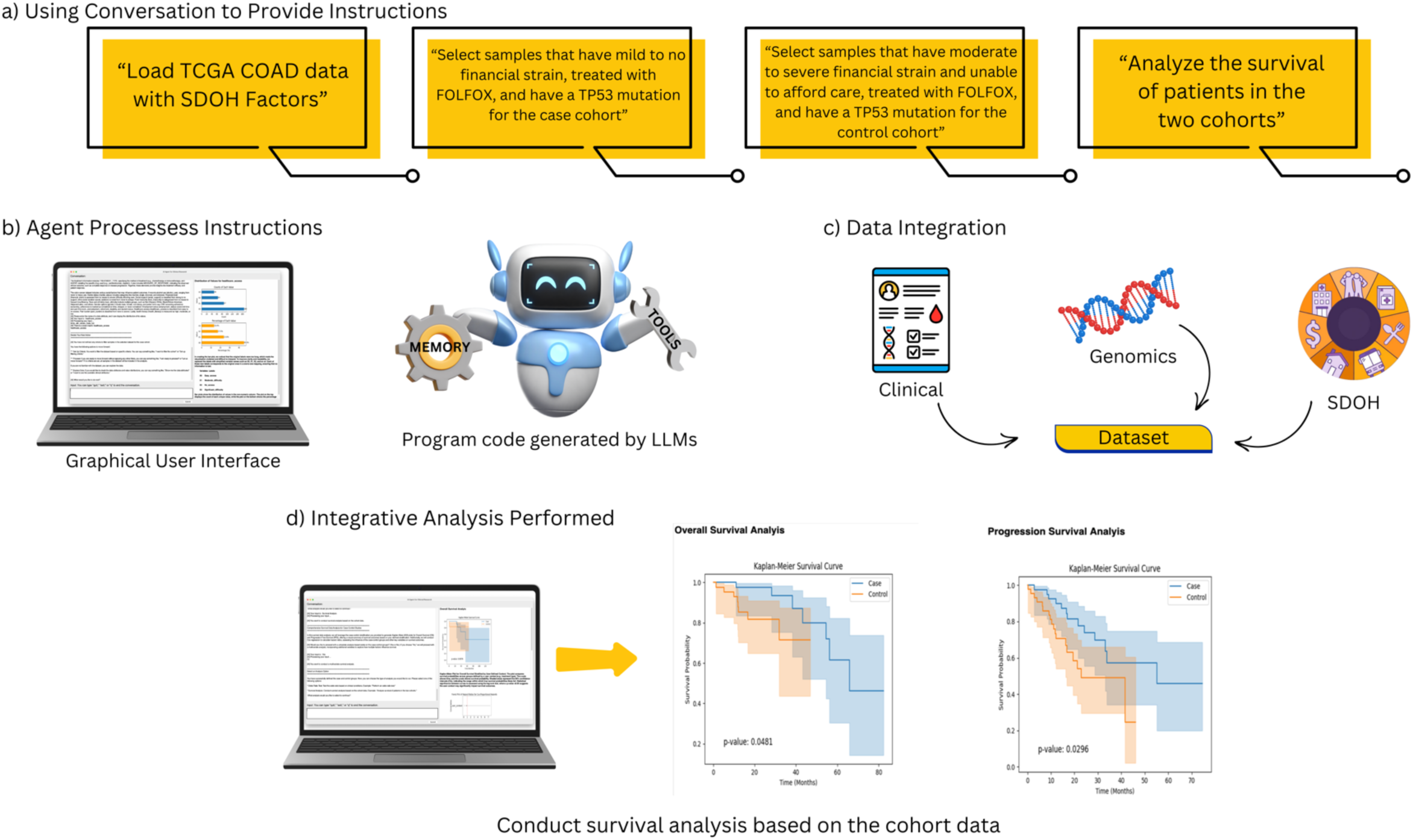
Overview of AI-HOPE-OM Workflow. This figure illustrates the workflow of the AI-HOPE-PM system for data analysis through a conversational interface. a) Using conversation to provide instructions, the AI agent receives natural language instructions from the user, such as loading a colorectal cancer (CRC) dataset, selecting case and control cohorts based on specified filtering criteria, and analyzing the survival of patients in the two cohorts. b) The agent processes the instructions via a graphical user interface (GUI) that leverages a large language model (LLM). The LLM generates the necessary program code to execute tasks, utilizing memory and reasoning tools to ensure accurate data extraction and manipulation. c) The system automates bioinformatics tasks while integrating clinical, genomic, and SDOH data. d) Integrative data analysis is performed with the option to either run a survival analysis or odds ratio test based on the defined case and control cohorts. The results are displayed for interpretation and further research.

Unlike traditional GUI tools, AI-HOPE-PM supports complex, user-defined queries such as: “Analyze FOLFOX-treated colorectal cancer (CRC) patients with TP53 mutations and varying levels of financial strain.” The system autonomously identifies relevant data, filters patient cohorts, integrates clinical treatment and genomic mutation data with socioeconomic context, and generates statistical visualizations, survival curves, and interpretative summaries (Figure 2). By enabling real-time, dynamic exploration of clinical-genomic-SDoH interactions, AI-HOPE-PM simplifies complex workflows and enhances the translational relevance of precision oncology research.

**Figure 2:**
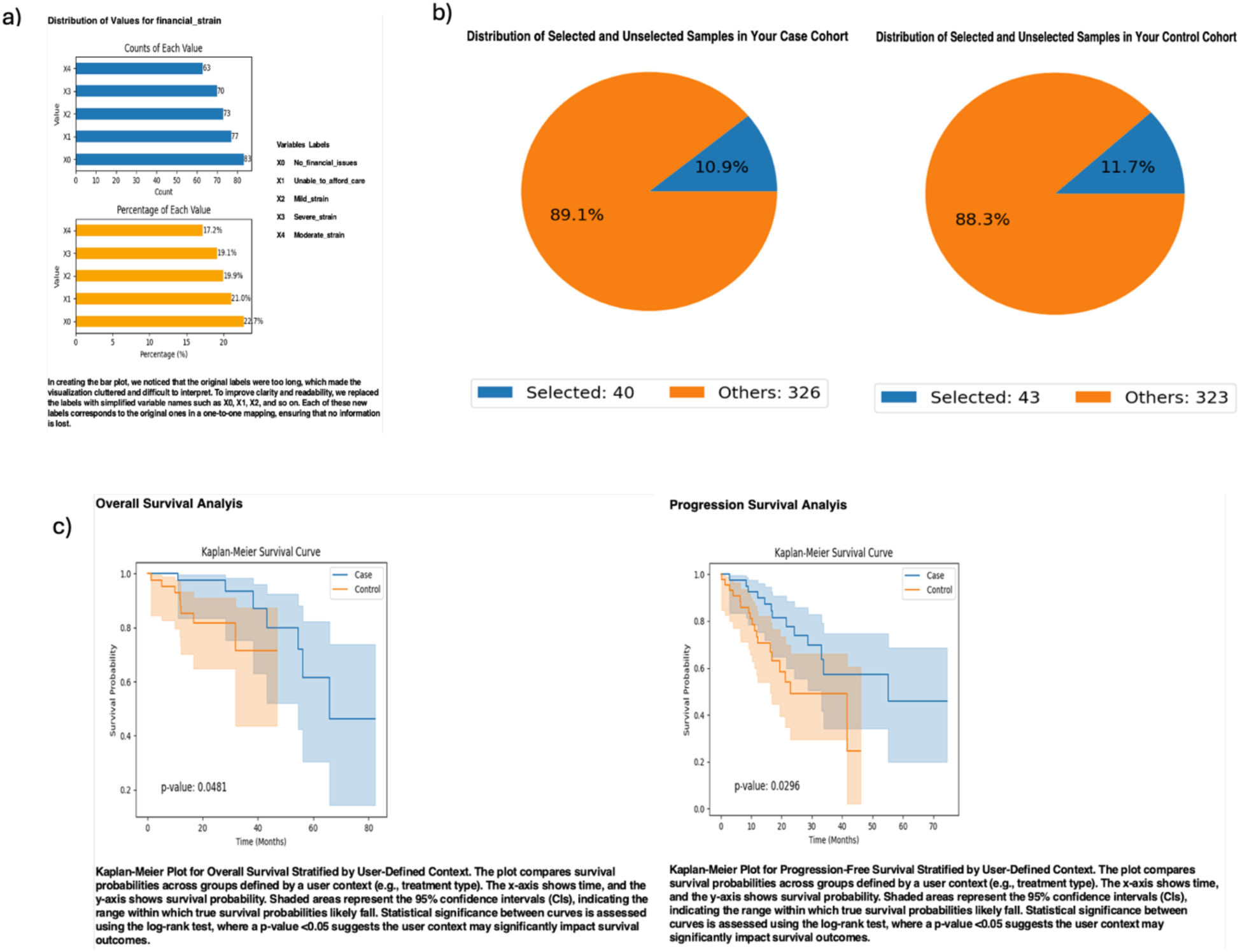
AI-HOPE-PM Analysis of FOLFOX-Treated CRC Patients with TP53 Mutations and Varying Levels of Financial Strain. This figure highlights the key steps in AI-HOPE-PM’s analysis of overall and progression-free survival among colorectal cancer (CRC) patients filtered into case and control cohorts. a) Selecting the COAD dataset with social determinants of health (SDOH) will provide the user with the ability to explore all the data attributes and value distributions. This bar chart illustrates the distribution of financial strain variables within the dataset. The blue bar chart shows the count of samples for each financial strain variable while the orange bar chart displays the corresponding percentage distribution. b) Defining the criteria to filter the samples in the selected dataset creates a case and control cohort to be used in the analysis. When the agent is provided with the defining criteria to refine the samples, pie charts are produced that illustrate the sample distribution among the case and control cohort. Out of 366 total samples, the case cohort is comprised of 40 samples (10.9%) that matched the filtering criteria of mild financial strain to no financial issues, treated with FOLFOX, and have a mutation in TP53. The control cohort consists of 43 samples (11.7%) that matched the same filtering criteria except for financial strain ranging from moderate to severe, including being unable to afford care. c) Once the case and control cohorts are successfully defined, the user can proceed and choose the type of analysis to run. Running a survival data analysis for case-control studies produces a Kaplan-Meyer survival curve which evaluates overall and progression-free survival. The overall (left) and progression-free (right) survival plots demonstrate that patients in the control cohort had significantly shorter survival compared to those in the case cohort, as indicated by the p-value of 0.0481 and 0.0296 respectively along with the confidence intervals.

To evaluate its performance, AI-HOPE-PM is being benchmarked against established tools such as cBioPortal and UCSC Xena [21]. The benchmarking involves assessing runtime efficiency, reproducibility, and usability. In contrast to tools that require step-by-step configuration, AI-HOPE-PM offers streamlined execution of advanced bioinformatics pipelines through LLM-guided user interaction, significantly lowering barriers to data exploration and hypothesis testing.

By bridging the gap between data complexity and user accessibility, AI-HOPE-PM offers a scalable, inclusive, and equitable AI framework for biomedical discovery. Its ability to integrate clinical, genomic, and SDoH variables addresses the long-standing need for tools that not only generate high-quality insights but also promote diversity and inclusiveness in biomedical research.

## Methods

### AI-HOPE-PM Architecture and Workflow

AI-HOPE-PM (Artificial Intelligence agent for High-Optimization and Precision mEdicine in Population Metrics) is a conversational artificial intelligence system that facilitates integrative biomedical data analysis through natural language interaction. The system leverages structured NLP, retrieval-augmented generation (RAG), and executable workflow translation to empower users to define complex case-control studies without requiring programming expertise. Once queries are submitted, the platform autonomously conducts cohort definition, statistical testing, and output generation in real time (Figure 1). The analytical process includes case selection, genomic and SDoH-based filtering, statistical analyses such as survival modeling and odds ratio testing, and automatic generation of visual and textual results.

### Data Integration and Preparation

AI-HOPE-PM operates on curated multi-modal datasets integrating clinical, genomic, and social determinants of health (SDoH) data. These datasets are sourced from publicly available repositories, including The Cancer Genome Atlas (TCGA), cBioPortal, and the AACR GENIE project, and are formatted into standardized tab-delimited files with associated metadata descriptors. Each dataset is accompanied by an index file outlining core variables such as overall survival, progression-free survival, treatment information, mutational status, demographic data, and SDoH indicators. The SDoH variables—including financial strain, food insecurity, health literacy, race/ethnicity, social support, and healthcare access—were selected through a comprehensive literature review and generated using simulated data via a Python script, which is included in the Materials section for reproducibility. The platform automates ingestion, formatting, and validation of all data files, ensuring analytical consistency and minimizing preprocessing burden.

### Natural Language Querying and Cohort Subsetting

Users interact with AI-HOPE-PM using plain-text questions or commands, which the system processes through a LLM and parses into structured database queries. The platform dynamically interprets filtering criteria, enabling sample stratification based on gene mutations, treatment regimens, clinical parameters, and SDoH variables. For instance, users may request to analyze CRC patients treated with FOLFOX who harbor TP53 mutations and differ by level of financial strain, or compare outcomes among patients with and without APC mutations under similar healthcare conditions. When faced with ambiguity or incomplete instructions, the system prompts users for clarification, enhancing accuracy and reducing risk of invalid analyses.

### Statistical and Bioinformatics Analysis

AI-HOPE-PM supports a broad range of analytical workflows. For categorical comparisons, the system performs prevalence estimation and odds ratio analysis using appropriate statistical tests to evaluate differences across defined patient subgroups.

For time-to-event data, Kaplan-Meier survival curves are generated with log-rank tests to assess significance, and Cox proportional hazards regression models are employed to quantify effect sizes across clinical, genomic, and SDoH variables. The platform also supports multivariable comparisons, allowing researchers to explore interactions between genetic alterations and behavioral, demographic, or social exposures in relation to clinical outcomes. All analyses are executed via an internal Python-based framework that adheres to established statistical conventions.

### Engineering Framework and Ontology Integration

Unlike direct prompting of LLaMA 3, AI-HOPE-PM incorporates an intermediate software layer that enforces query structure, biomedical ontology alignment, and analysis integrity. Structured prompts are engineered to ensure output adherence to standard practices in genomics and epidemiology. Retrieval-augmented generation enables live referencing of validated public data from TCGA and cBioPortal, while custom modules translate queries into executable bioinformatics scripts. To mitigate hallucination risk and ensure reproducibility, AI-HOPE-PM applies structured output validation and incorporates biomedical ontologies to verify entity recognition and relationship mapping within each query.

### Computational Requirements and Scalability

The platform is optimized to run on CPU-or GPU-enabled environments, including cloud-based infrastructures such as AWS, Azure, and Google Cloud. Minimum system requirements include an Intel Xeon or comparable processor with four or more cores, 16 GB of RAM, and 1 TB of SSD storage for handling large genomic datasets. While CPU deployment is sufficient for standard analysis, GPU acceleration using hardware such as the NVIDIA A100 is recommended for large-scale queries, particularly those involving extensive natural language interpretation or deep learning-based prediction models. AI-HOPE-PM supports parallel execution and distributed computing for population-level data analysis, ensuring scalable performance for high-throughput research needs.

### Interpretability and Evaluation of Language Processing

To evaluate its ability to translate natural language into bioinformatics workflows, AI-HOPE-PM was tested against 100 diverse biomedical research queries spanning genomics, clinical data, and social determinant themes. The system achieved a natural language interpretation accuracy of 92.5%, outperforming other AI-based tools, with 99.1% accuracy for simple queries and 88.4% for complex, multivariable prompts.

Errors primarily arose in queries lacking explicit references, which were mitigated through AI-HOPE-PM’s built-in context-aware prompting for clarification.

### Benchmarking Workflow Efficiency Across Platforms

To evaluate computational efficiency and usability, we conducted a benchmark test comparing AI-HOPE-PM with widely used platforms cBioPortal and UCSC Xena. The test focused on measuring the time required to open each application, select a relevant cancer dataset, and filter based on a single data attribute. These tasks were performed by experienced biomedical informatics researchers who are frequent users of cBioPortal and UCSC Xena, ensuring a fair and skill-appropriate comparison. Each platform was accessed under similar computing conditions, and task execution times were recorded in seconds using a standardized stopwatch protocol. This experiment aimed to simulate real-world usage and assess the responsiveness and automation advantages offered by AI-HOPE-PM relative to established tools.

### Automated Reporting and Benchmarking

After executing a query, AI-HOPE-PM generates structured reports that include survival curves, odds ratios, statistical summaries, and narrative interpretations based on validated literature. Reports also include visualizations such as bar charts, Kaplan-Meier plots, and forest plots to facilitate dissemination and interpretation. The system is being benchmarked against cBioPortal and UCSC Xena to evaluate performance, runtime, and reproducibility. Direct comparisons with other generative AI systems such as CellAgent and AutoBA were deemed non-comparable due to AI-HOPE-PM’s unique integration of SDoH data. A formal usability study is ongoing to assess accessibility and efficiency relative to GUI-based analysis pipelines.

## Results

By converting natural language instructions into executable bioinformatics workflows, AI-HOPE-PM enabled seamless integration and analysis of clinical, genomic, and social determinants of health (SDoH) data within colorectal cancer (CRC) datasets. The platform’s ability to interpret user queries and automate complex analyses demonstrated its effectiveness in supporting multi-dimensional, translational cancer research. Through its intuitive conversational interface, the system dynamically classified patient samples into case and control cohorts based on user-defined criteria. These criteria encompassed gene mutation status, treatment regimens, SDoH attributes, and demographic variables, facilitating highly customizable stratifications. The system autonomously performed statistical analyses—including prevalence estimation, odds ratio tests, and survival modeling—and generated comprehensive visualizations and interpretable reports.

In a prominent use case, AI-HOPE-PM analyzed data from the TCGA COAD dataset to investigate how financial strain affects outcomes among FOLFOX-treated CRC patients with TP53 mutations (Fig. 2). Based on user-defined filters, the system identified a case cohort of 40 patients (10.9%) with no or mild financial strain and a control cohort of 43 patients (11.7%) with moderate to severe financial strain, including individuals unable to afford care. Kaplan-Meier analysis revealed significantly poorer overall and progression-free survival in the control group, with p-values of 0.0481 and 0.0296, respectively. These findings illustrate AI-HOPE-PM’s capacity to integrate genomic, treatment, and SDoH data to uncover clinically relevant disparities in survival outcomes.

Another case study explored the impact of APC mutation status among CRC patients treated with FOLFOX and reporting easy access to healthcare (Fig. 3). AI-HOPE-PM filtered the SocialFactor_COAD dataset to define 40 APC-mutated patients and 12 wild-type APC patients. Kaplan-Meier analysis revealed significantly worse progression-free survival among APC wild-type patients (p = 0.0233). Odds ratio analysis showed lower, though not statistically significant, Hispanic/Latino representation in the control group (OR = 0.529; CI: [0.11, 2.541]), reinforcing the need to consider ancestral background and access to care in precision oncology.

**Figure 3:**
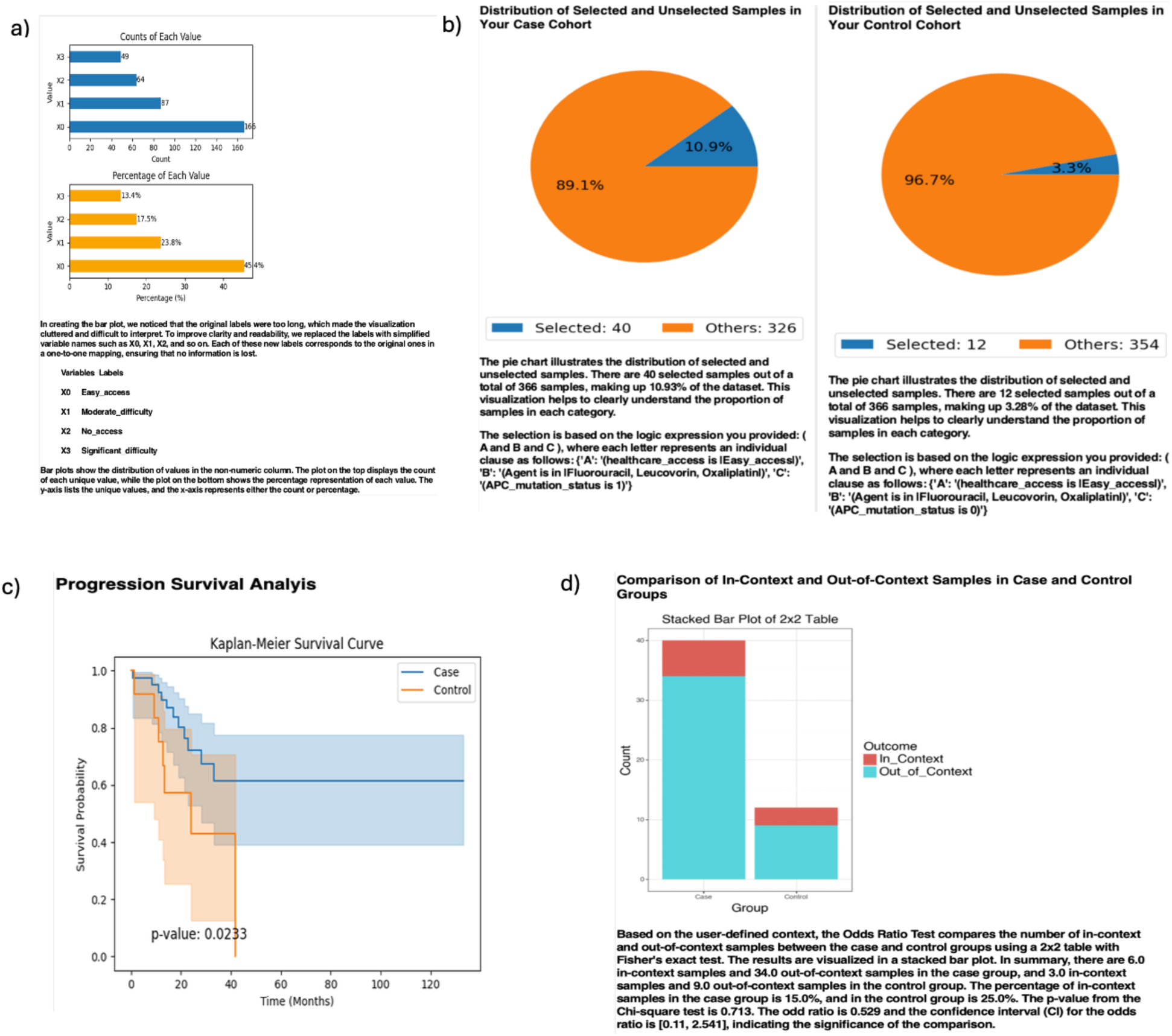
AI-HOPE-PM Analysis of CRC Patients with and without APC mutations that have Easy Access to Healthcare and Treated with FOLFOX. This figure highlights the key steps in AI-HOPE-PM’s analysis of colorectal cancer (CRC) patients who have easy access to healthcare, have been treated with FOLFOX, and either have or don’t have APC mutations. a) Selecting the COAD dataset with social determinants of health (SDOH) will provide the user with the ability to explore all the data attributes and value distributions. This bar chart illustrates the distribution of healthcare access variables within the dataset. The blue bar chart shows the count of samples for each healthcare access variable while the orange bar chart displays the corresponding percentage distribution. b) Defining the criteria to filter the samples in the selected dataset creates a case and control cohort to be used in the analysis. When the agent is provided with the defining criteria to refine the samples, pie charts are produced that illustrate the sample distribution among the case and control cohort. Out of 366 total samples, the case cohort is comprised of 40 samples (10.9%) that matched the filtering criteria of easy healthcare access, treated with FOLFOX, and APC mutated. The control cohort only has 12 samples (3.3%) that matched the same filtering criteria except these samples did not have APC mutated. c) Once the case and control cohorts are successfully defined, the user can proceed and choose the type of analysis to run. Running a survival data analysis for case-control studies produces a Kaplan-Meyer survival curve which evaluates overall and progression-free survival. The plot demonstrates that patients in the control cohort had significantly poorer progression-free survival compared to those in the case cohort, as indicated by the p-value and confidence intervals. d) Another analysis option is the odds ratio test, in which the user must define the context to compare between the case and control groups. The context used for this analysis was ethnicity being Hispanic-Latino which resulted in 6 in-context samples (15%) and 34 out-of-context samples in the case group and 3 in-context samples (15%) and 9 out-of-context samples in the control group. The odd ratio was 0.529 and the confidence interval for the odds ratio was [0.11, 2.541].

In a third application, the system examined early-onset CRC patients (age <50) to evaluate the role of social support on survival among those treated with FOLFOX (Fig. S1). The case cohort comprised 17 patients with strong or moderate social support, while the control cohort included 14 patients with limited or no support. Although the overall survival difference was not statistically significant (p = 0.0741), progression-free survival was significantly poorer in the control group (p = 0.0220). An accompanying odds ratio test evaluating TP53 mutation status yielded an OR of 0.706 (CI: [0.208, 2.396]), suggesting a possible link between psychosocial factors and mutational context in younger patients.

In another analysis focused on food insecurity, AI-HOPE-PM investigated survival disparities among CRC patients with APC mutations (Fig. S2). The system identified 80 patients (21.9%) with no reported food insecurity and 186 patients (50.8%) with moderate to severe food insecurity. Kaplan-Meier analysis revealed that food-insecure patients experienced significantly worse progression-free survival (p = 0.0162). Odds ratio testing further indicated a lower likelihood of receiving chemotherapy among food-insecure patients (OR = 0.356; CI: [0.136, 1.186]), underscoring the role of socioeconomic burden in modulating treatment access and outcomes.

Sex-based disparities were explored in a separate study focusing on CRC patients with limited health literacy treated with FOLFOX (Fig. S3). AI-HOPE-PM created a case cohort of 33 female and a control cohort of 41 male patients. Odds ratio testing using KRAS mutation status as the context revealed that 30.3% of the case group and 56.1% of the control group had KRAS mutations, with an odds ratio of 0.503 (CI: [0.192, 1.319], p = 0.244). Though not statistically significant, these results suggest potential sex-based differences in mutation prevalence under constrained health literacy conditions.

AI-HOPE-PM also facilitated analyses of non-genomic SDoH influences. One study revealed that moderate to severe financial strain significantly reduced CRC screening adherence, corroborating known links between economic hardship (Fig. S4) and healthcare access (Fig. S5). Another highlighted that patients reporting low social support or isolation had elevated rates of treatment discontinuation and significantly worse survival, aligning with psychosocial oncology literature [22]. The system further identified racial and ethnic disparities in progression-free survival, with non-Hispanic White patients faring better than their Black and Hispanic counterparts, even after adjusting for treatment type and disease stage. These results underscore the value of incorporating SDoH into precision medicine frameworks.

AI-HOPE-PM demonstrated high computational efficiency, executing high-dimensional case-control studies involving over 10,000 patient records in under one minute. In a benchmark comparison, the platform required only 28.02 seconds to open the application, select a database, and filter a single data attribute—significantly faster than cBioPortal (58.01 seconds) and UCSC Xena (46.06 seconds). By automating the ingestion, filtering, analysis, and reporting stages, AI-HOPE-PM substantially reduced manual burden and turnaround time compared to conventional bioinformatics tools. This performance underscores its value as a scalable AI platform capable of delivering real-time, integrative data analysis to support precision oncology and health equity research.

## Discussion

AI-HOPE-PM marks a significant leap in precision medicine by unifying clinical, genomic, and social determinants of health (SDoH) data within a single, conversational AI platform. Rather than relying on traditional tools, the system autonomously interprets complex, user-defined natural language queries, converts them into robust bioinformatics workflows, and performs multi-layered statistical analyses—all while delivering interpretable outputs. In doing so, it overcomes key limitations of conventional genomic analysis platforms.

A key strength of AI-HOPE-PM lies in its flexibility and interactivity, allowing users to define nuanced case-control stratifications that consider both biological and non-biological variables. For example, in the case of FOLFOX-treated CRC patients with TP53 mutations, AI-HOPE-PM revealed a clear association between financial strain and survival outcomes. This type of multi-dimensional insight, integrating treatment history, gene mutation, and socioeconomic burden, would be difficult to achieve using static platforms such as cBioPortal or UCSC Xena, which often lack support for SDoH variables and require users to perform multi-step manual filtering.

Importantly, AI-HOPE-PM extends precision medicine beyond the genomic level by incorporating SDoH into its analytic framework. The observed associations between food insecurity and progression-free survival, as well as the impacts of social support on early-onset CRC outcomes, reflect the growing recognition that social and behavioral factors critically influence cancer trajectories. These results emphasize that genomic discoveries must be contextualized within broader environmental and social landscapes to ensure that treatment strategies are equitable and population-relevant.

AI-HOPE-PM’s application of advanced natural language processing, powered by LLaMA 3 and supported by retrieval-augmented generation (RAG) and biomedical ontology constraints, also enhances its reliability and reproducibility. Unlike generic AI chatbots, AI-HOPE-PM dynamically translates user prompts into executable code aligned with established bioinformatics methodologies, ensuring statistical rigor while minimizing AI hallucinations. The platform’s reported 92.5% accuracy in interpreting user queries underscores its potential to democratize data-driven research and bridge the technical gap for clinicians and public health researchers.

AI-HOPE-PM’s superior computational performance illustrates its potential to transform how researchers and clinicians interact with complex biomedical datasets. The system completed high-dimensional case-control analyses involving over 10,000 patient records in under one minute, demonstrating exceptional scalability for real-time translational research. In benchmarking tests, AI-HOPE-PM performed essential tasks—such as launching the platform, selecting a dataset, and filtering a single attribute—in around half the time required by established tools like cBioPortal and UCSC Xena (Table S1). This efficiency reflects its streamlined architecture and fully automated workflow, which together reduce the manual workload and eliminate common bottlenecks in traditional bioinformatics pipelines. By accelerating analytic tasks and minimizing the need for technical expertise, AI-HOPE-PM enhances both productivity and accessibility, reinforcing its potential as a powerful, equitable platform for precision oncology and health equity research.

While these results are promising, continued benchmarking of AI-HOPE-PM against state-of-the-art platforms such as CellAgent [15] and AutoBA [16] is essential to fully assess its comparative advantages in multi-omics [23], spatial biology [24] and single cell genomic [25–27] analysis. Additionally, further optimization of the natural language pipeline and integration with federated learning models could enhance its applicability in clinical environments that demand secure and continuous data updates.

### Conclusion

AI-HOPE-PM offers a significant advancement in precision medicine by seamlessly integrating clinical, genomic, and social determinants of health (SDoH) data into a single, AI-driven analytic platform. Leveraging a natural language interface and automated bioinformatics workflows, the system removes technical barriers, allowing both researchers and clinicians to perform complex, multi-modal analyses without coding expertise. This functionality enables nuanced cohort stratification based on genomic profiles, treatment regimens, and SDoH variables, fostering a more comprehensive understanding of cancer biology and health disparities. By delivering analyses that are rapid, reproducible, and easily interpretable, AI-HOPE-PM not only accelerates biomarker discovery but also supports equitable and evidence-based decision-making in oncology. With its scalability and adaptability, the platform is well-positioned to drive forward both innovation in cancer research and progress in inclusive cancer care.

## Data Availability

All data used in the present study is publicly available at https://www.cbioportal.org/, https://genie.cbioportal.org and https://xena.ucsc.edu. Additional data can be provided upon reasonable request to the authors.

---

**Figure S1:**
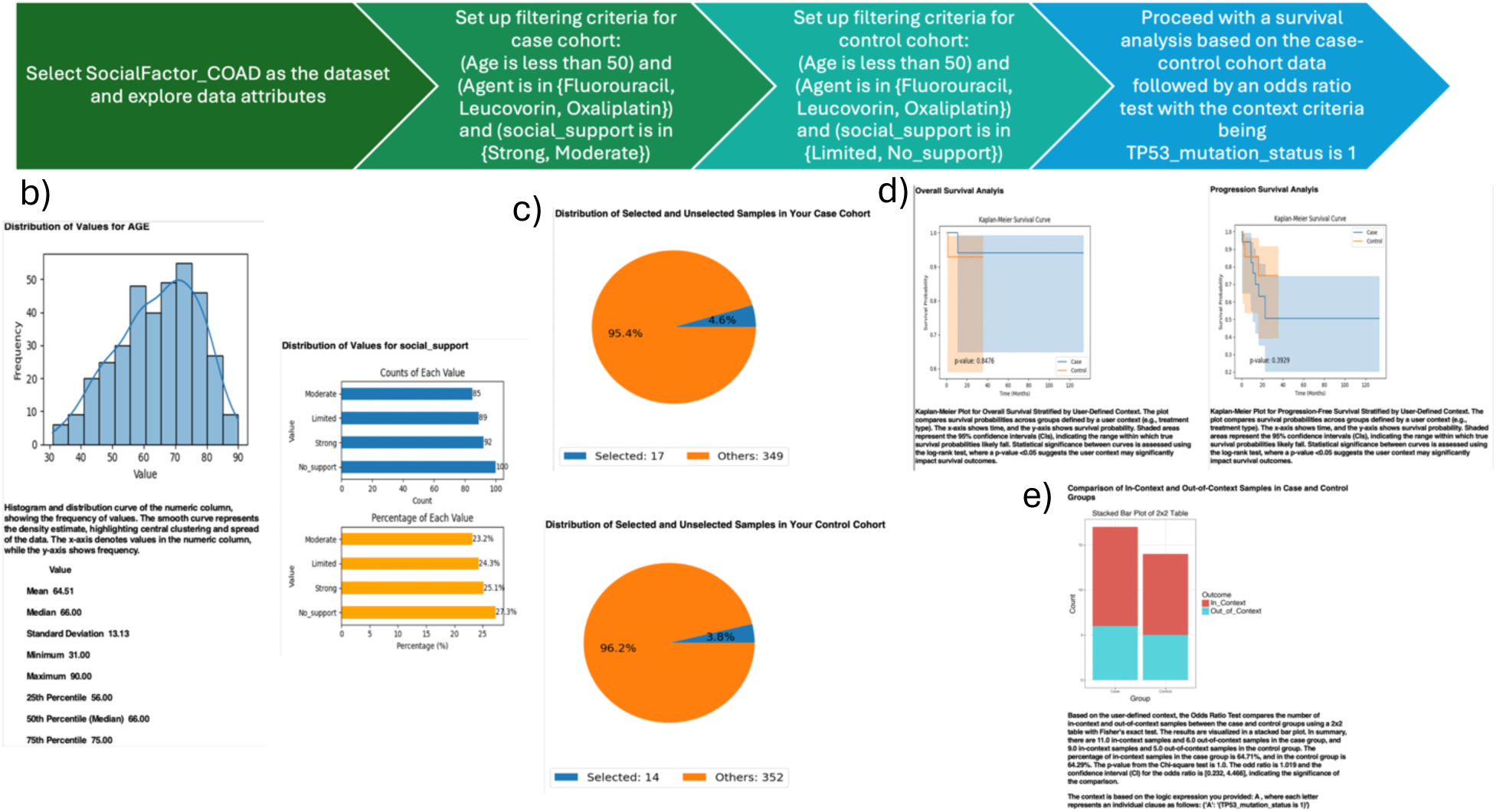
AI-HOPE-PM Analysis of Early-Onset CRC Patients Treated with FOLFOX and Varying Levels of Social Support. a) The user begins by selecting the COAD dataset enriched with social determinants of health (SDOH) attributes. This step enables exploration of patient-level variables such as age, treatment type, mutation status, and social support levels. The histograms and bar plots on the left display the distribution of these attributes within the dataset. b) The user sets the filtering criteria to define the case cohort, which includes patients under age 50, treated with FOLFOX (5-Fluorouracil, Leucovorin, Oxaliplatin), and classified as having strong or moderate social support. This results in a cohort of 17 patients. The orange and blue pie chart shows the proportion of selected versus total samples. c) In parallel, the control cohort is defined using the same criteria except for social support, selecting patients with limited or no support, resulting in 14 patients. The corresponding pie chart visualizes the distribution of selected control samples. d) Once both cohorts are created, a Kaplan-Meier survival analysis is performed. The survival curves for overall and progression-free survival indicate a significant diVerence between the groups. While the diVerence in overall survival is not statistically significant (p = 0.0741), progression-free survival is significantly poorer in the control group (p = 0.0220), suggesting that lower levels of social support may be linked to worse clinical outcomes. e) An additional odds ratio analysis was performed using TP53 mutation status as the defining context. In this analysis, TP53 mutation status was evaluated between the two groups, resulting in an odds ratio of 0.706 with a confidence interval of [0.208, 2.396].

**Figure S2:**
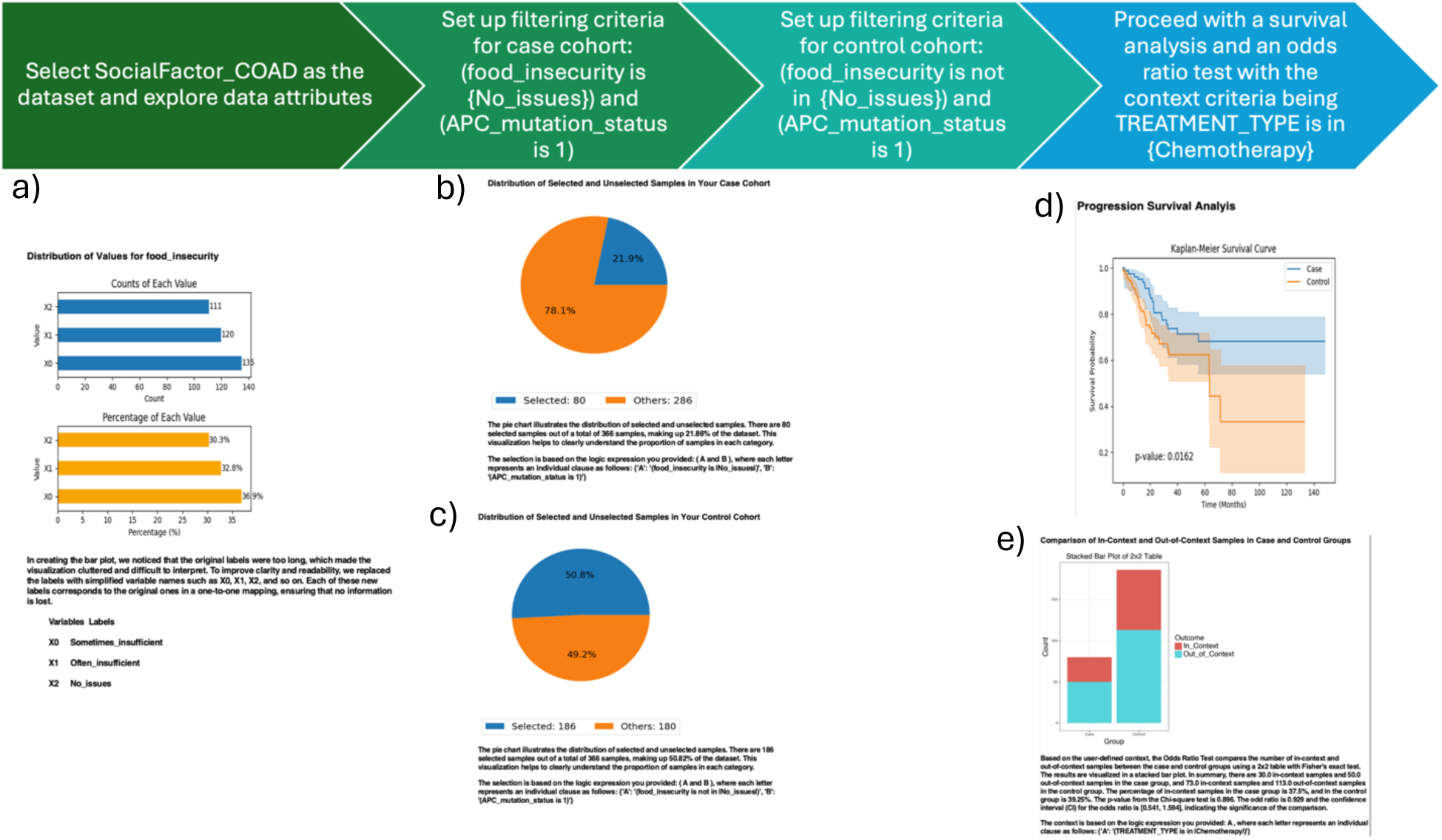
AI-HOPE-PM Analysis of CRC Patients with and without Chemotherapy Treatment, Food Security, and APC Mutations. This figure demonstrates the AI-HOPE-PM platform’s analysis of colorectal cancer (CRC) patients with a focus on food insecurity, APC mutation status, and treatment type. The study compares progression-free survival and treatment disparities among patients with similar genomic and social contexts. a) The analysis begins by selecting the COAD dataset integrated with social determinants of health (SDOH). Data attributes such as food insecurity, treatment type, and APC mutation status are visualized using histograms and bar plots. The top-left bar chart displays the distribution of food insecurity across the dataset, while the orange and blue charts summarize mutation and treatment type proportions. b) The case cohort is created by applying filters for patients who report no food insecurity and are APC-mutated (mutation status = 1). This results in 245 samples. A pie chart illustrates the proportion of these case samples in relation to the overall dataset. c) Similarly, the control cohort is defined using the same food insecurity and APC mutation criteria. This results in 206 samples for the control group. A corresponding pie chart visualizes the breakdown of selected samples for this cohort. d) With the case and control cohorts defined, a progression-free survival analysis is conducted. The Kaplan-Meier curve shows diVerences in outcomes between the two groups, stratified by treatment type, specifically chemotherapy. Although full statistical values (e.g., p-values) are not specified in the visual, the separation in curves indicates diVerential survival based on treatment in the context of food security and mutation status. e) An odds ratio analysis is also performed using TREATMENT_TYPE (chemotherapy vs. non-chemotherapy) as the defining context between case and control groups. The results, shown in the bar plot, suggest a distribution diVerence in chemotherapy treatment exposure. This analysis allows researchers to examine how social context (e.g., food insecurity) and genomic status intersect with treatment access and outcomes.

**Figure S3:**
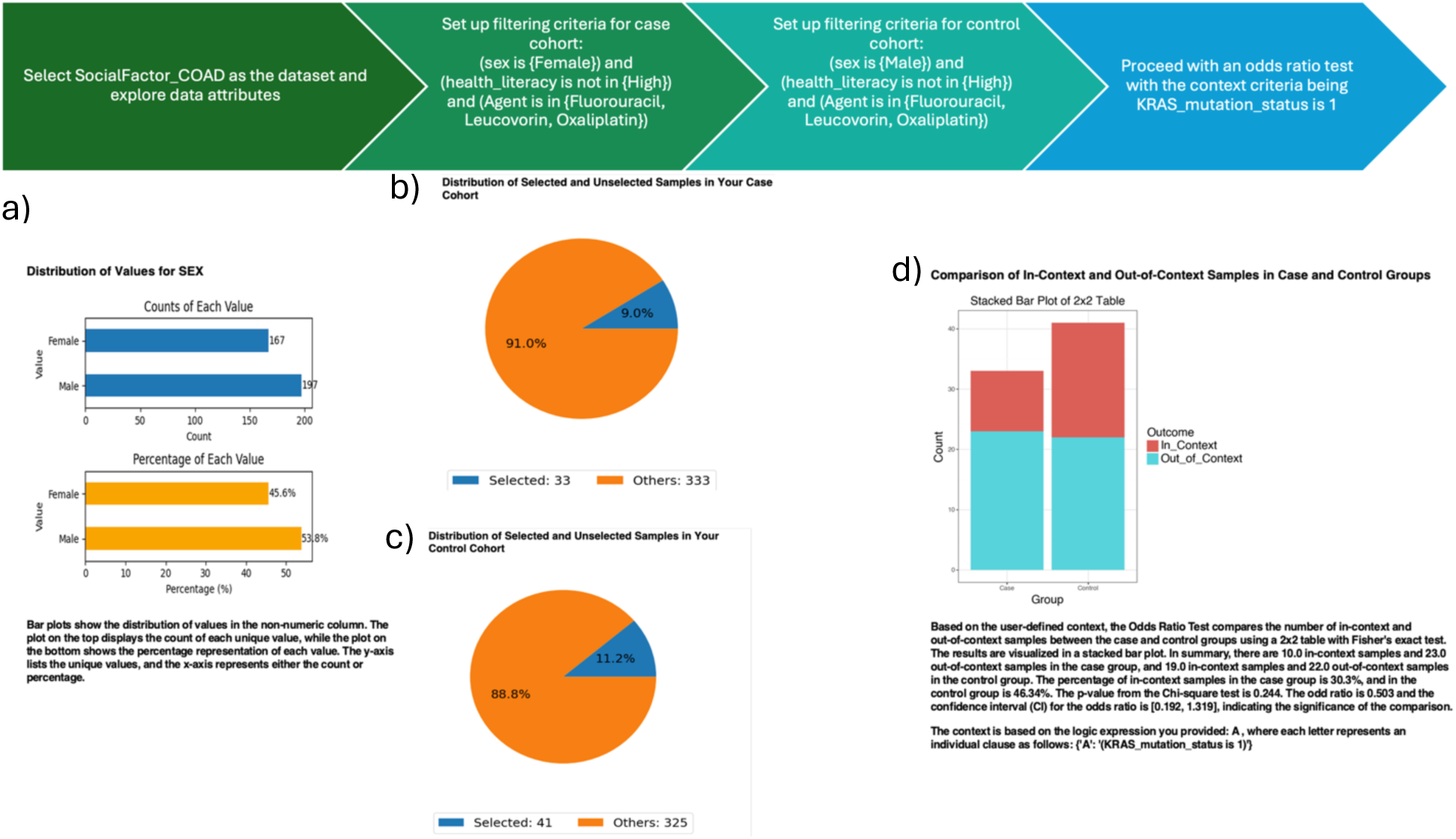
AI-HOPE-PM Analysis of CRC Patients with KRAS Mutations in the Context of Insurance Coverage and Tumor Stage. This figure presents an AI-HOPE-PM analysis of colorectal cancer (CRC) patients within the COAD dataset, focusing on how insurance status, tumor stage, and KRAS mutation status intersect to influence patient stratification and potential disparities in care. a) The user first selects the COAD dataset enriched with social determinants of health (SDOH) and genomic variables. The bar charts display the distribution of insurance status within the dataset. The top panel shows the count of patients by insurance category, while the bottom panel presents the corresponding percentages. This provides an overview of how patients are distributed based on access to financial resources for care. b) The case cohort is defined by filtering for patients who are insured, have KRAS mutations (mutation_status = 1), are in Stage I or Stage II, and received Leucovorin chemotherapy treatment. The resulting pie chart indicates that 31 samples met these criteria out of a total of 373 available samples. c) The control cohort is generated using the same filtering criteria, except selecting uninsured patients. A second pie chart illustrates that this cohort includes 30 samples. The visual comparison helps highlight how insurance coverage may influence treatment access and patient stratification despite identical clinical and molecular parameters. d) An odds ratio test is then conducted to explore diVerences in treatment exposure between the insured and uninsured groups, using KRAS mutation status as the defining context. The stacked bar chart visualizes the proportion of in-context (KRAS-mutated) and out-of-context samples in each group. The results suggest a modest diVerence in KRAS mutation representation between insured and uninsured cohorts, which may warrant further exploration of how financial and genomic factors interact to shape treatment decisions and patient outcomes.

**Figure S4.**
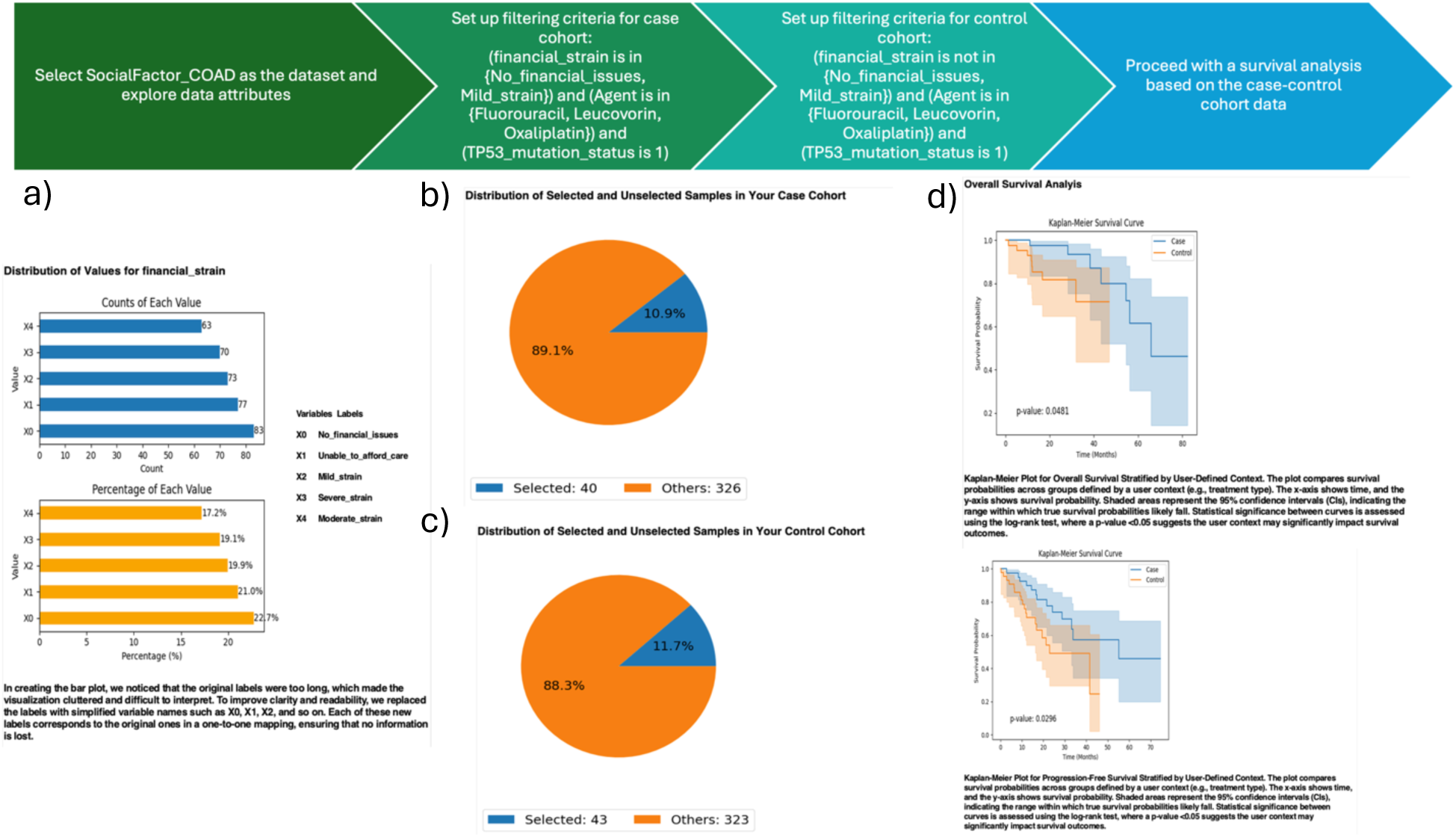
AI-HOPE-PM Analysis of Survival Outcomes in CRC Patients with DiVerent Insurance and Treatment Profiles. This figure outlines an AI-HOPE-PM case-control analysis of colorectal cancer (CRC) patients using the COAD dataset, focusing on the impact of insurance status, treatment exposure, and social determinants of health (SDOH) on overall survival outcomes. a) The analysis begins with the selection of the COAD dataset enriched with SDOH attributes. Bar charts on the left show the distribution of insurance types among the cohort. The top chart displays the absolute count of patients per insurance category, while the bottom chart shows the corresponding percentage distribution. These visualizations oVer context for the socioeconomic landscape of the dataset. b) The case cohort is defined by selecting insured patients with the following characteristics: Stage IV diagnosis, Hispanic/Latino ethnicity, treated with FOLFOX, and who received care at a Community Oncology Practice (COP). This cohort contains 41 patients, as depicted by the pie chart illustrating the proportion of selected samples within the total case pool. c) The control cohort is created using the same criteria, except for patients who are uninsured. This results in 22 patients, visualized in a separate pie chart. The comparison highlights disparities in treatment access and healthcare utilization between insured and uninsured groups under otherwise similar clinical and demographic conditions. d) With the cohorts defined, the system proceeds with a Kaplan-Meier survival analysis. Two plots illustrate overall survival diVerences between the case and control groups, stratified by insurance and treatment status. The survival curves show a noticeable separation, suggesting worse survival outcomes for uninsured patients, although the p-values and confidence intervals would determine statistical significance. These results underscore the impact of insurance coverage on survival, even when controlling for clinical and molecular variables.

**Figure S5.**
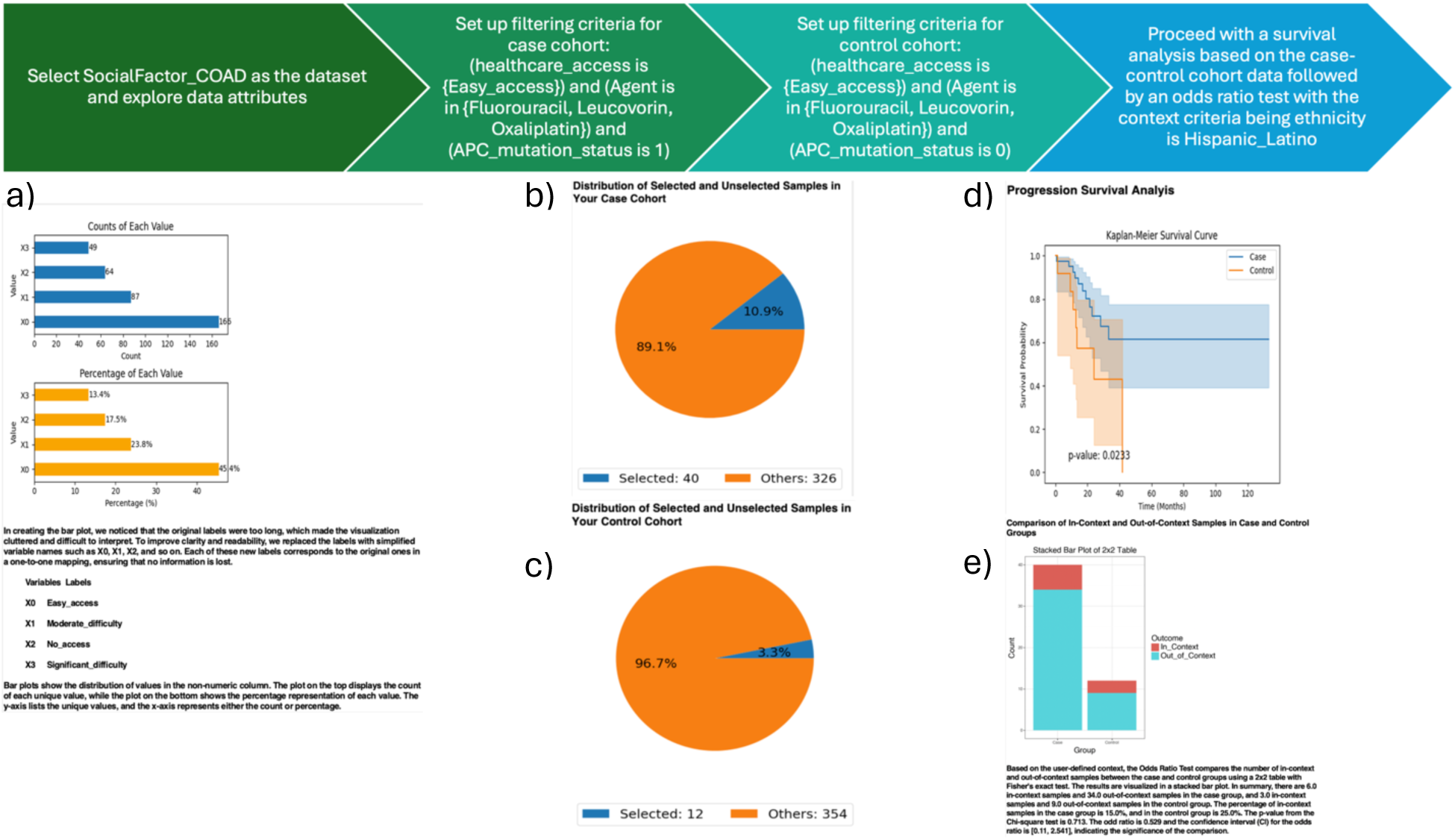
AI-HOPE-PM Stratification of CRC Patients by Healthcare Access, APC Mutation, and Ethnicity for Survival and Treatment Disparity Analysis. This figure highlights the AI-HOPE-PM platform’s ability to stratify colorectal cancer (CRC) patients using integrative filters based on healthcare access, genomic features, and ethnicity to study treatment disparities and survival outcomes. The platform enables a structured, stepwise case-control analysis within the SocialFactors_COAD dataset. a) The analysis begins by selecting the COAD dataset, enriched with social determinants of health (SDoH) data, and visualizing variables such as APC mutation status and healthcare access. Bar plots show the frequency of APC mutations and the distribution of healthcare access categories within the dataset. b) A case cohort is generated by applying filters for patients with limited healthcare access, specific treatment types (Agents in Fluorouracil, Leucovorin, Oxaliplatin), and APC mutation status = 1. A pie chart displays that 326 samples meet these criteria, showing their proportion relative to the full dataset. c) A control cohort is created using the same filtering criteria for healthcare access and treatment but with APC mutation status = 0. The resulting pie chart shows 354 control samples and their proportional representation. d) A survival analysis is performed using the case-control structure, stratifying patients based on chemotherapy treatment status. The Kaplan-Meier plot presents progression-free survival diVerences between groups, with a noticeable separation in curves, particularly for Hispanic/Latino patients. e) Finally, an odds ratio analysis is conducted to assess treatment disparities based on chemotherapy exposure across the stratified cohorts. A bar plot illustrates diVerences in treatment distribution, indicating how access to care and genomic context may influence treatment receipt and outcomes.

**Table S1.**
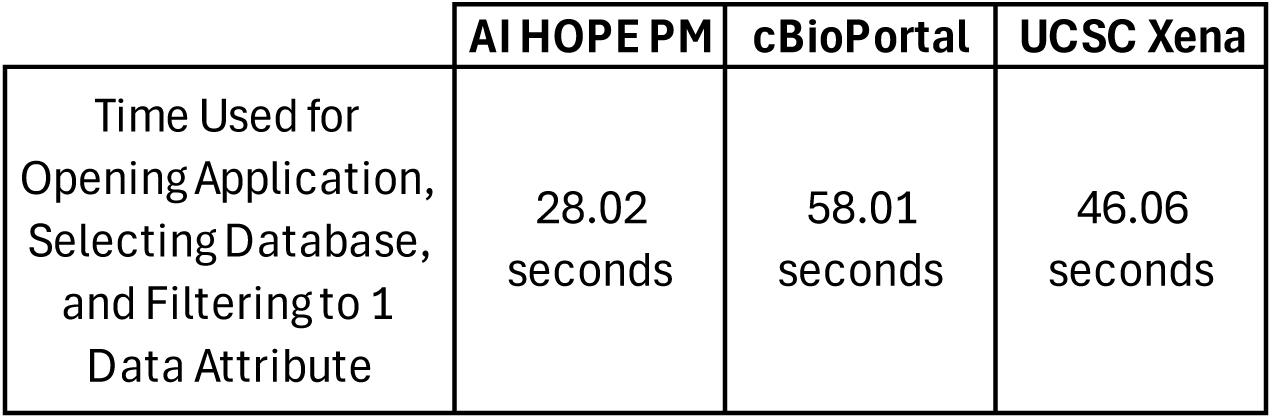
Comparative Timing Analysis for Executing Basic Data Query Tasks Across Platforms. This table summarizes the time (in seconds) required to complete a standardized task—opening the application, selecting a dataset, and filtering by one data attribute—across three platforms (AI-HOPE-PM, cBioPortal, UCSC Xena).

